# Evaluating the effectiveness of the smartphone app, Drink Less, compared with the NHS alcohol advice webpage, for the reduction of alcohol consumption among hazardous and harmful adult drinkers in the UK at six-month follow-up: protocol for a randomised controlled trial

**DOI:** 10.1101/2020.07.01.20144261

**Authors:** Claire Garnett, Melissa Oldham, Colin Angus, Emma Beard, Robyn Burton, Matt Field, Felix Greaves, Matthew Hickman, Eileen Kaner, Gemma Loebenberg, Susan Michie, Marcus Munafò, Elena Pizzo, Jamie Brown

## Abstract

**Background and Aims:** Digital interventions are effective for reducing alcohol consumption but evidence is limited regarding smartphone apps. *Drink Less* is a theory- and evidence-informed app to help people reduce their alcohol consumption that has been refined in terms of its content and design for usability across the socio-demographic spectrum. We aim to evaluate the effectiveness and cost-effectiveness of recommending *Drink Less* at reducing alcohol consumption compared with usual digital care.

**Design:** Two-arm individually randomised controlled trial.

**Setting:** Online trial in the UK.

**Participants:** Hazardous or harmful drinkers (Alcohol Use Disorders Identification Test score >=8) aged 18+, and want to drink less alcohol (n=5,562). Participants will be recruited from July 2020 to May 2022 using multiple strategies with a focus on remote digital methods.

**Intervention and comparator:** Participants will be randomised to receive either an email recommending that they use *Drink Less* (intervention) or view the NHS webpage on alcohol advice (comparator).

**Measurements:** The primary outcome is change in self-reported weekly alcohol consumption between baseline and 6-month follow-up. Secondary outcomes include the proportion of hazardous drinkers; alcohol-related problems and injury; health-related quality of life, and use of health services assessed at 6-month follow-up. Effectiveness will be examined with one-way ANCOVAs, adjusting for baseline alcohol consumption, and using an intention-to-treat approach. A mixed-methods process evaluation will assess engagement, acceptability and mechanism of action. Economic evaluations will be conducted using both a short- and longer-term time horizon.

**Comments:** This study will establish the effectiveness and cost-effectiveness of the *Drink Less* app at reducing alcohol consumption among hazardous and harmful adult drinkers and will be the first RCT of an alcohol reduction app for the general population in the UK. This study will inform the decision on whether it is worth investing resources in large-scale implementation.

## Introduction

Hazardous and harmful alcohol consumption is a major public health concern and contributes to health inequalities with the most deprived groups suffering the most harm from alcohol [1]. Fewer than 7% of hazardous and harmful drinkers receive face-to-face interventions in primary care to support alcohol reduction [2] with key barriers to the delivery of these interventions by practitioners being lack of time and low confidence about discussing alcohol with patients [3,4]. Digital interventions, such as websites and smartphone apps, may be effective for reducing alcohol consumption [5], and may overcome barriers to delivery of face-to-face interventions as they potentially have a broad reach and relatively low implementation costs (once developed), so can be delivered at scale [6]. As digital technologies become more integrated into everyday life and widely used, digital interventions can have a large positive impact on public health and wellbeing at a population level. Smartphone apps are a promising mode of intervention delivery because smartphones have become increasingly affordable to end users and prevalent among the UK population [7]. However, most digital alcohol interventions that have been evaluated are web-based and there is little evidence on the effectiveness of apps. The few trials of apps have been based in other countries and usually with younger adults [8–11]. Therefore, there is an urgent need for a robust evaluation of an evidence- and theory-informed alcohol reduction app, which, if effective, could be widely recommended to drinkers in the UK. The *Drink Less* app was designed to help people reduce their alcohol consumption, and has been developed and refined using a systematic and iterative process [12,13]. The app is ready for a definitive evaluation to establish whether recommending it to people is more effective than usual digital care for the reduction of alcohol consumption.

### What is already known

Digital interventions – primarily web-based – may reduce alcohol consumption, with an average reduction of 23g of alcohol (2.9 UK units) per week compared with participants in the control group [5]. In this Cochrane review of 42 RCTs, only one of the digital interventions used a smartphone app, and this RCT was conducted in Sweden amongst university students [8]. Updates of this review found a further three studies that used smartphone apps: one in Sweden amongst university students [9], one for the general population in Canada [10] and another for young adults in Australia [11]. Therefore, despite the availability of hundreds of alcohol-related apps, none have been evaluated in a RCT among the general population of adults in the UK. The majority have also been developed without reference to scientific evidence or theory [14]. The lack of evidence highlights the necessity of a robust and pragmatic evaluation of an evidence- and theory-informed alcohol reduction app, which, if effective, could be widely recommended.

### The Drink Less smartphone app

*Drink Less* is one of the most popular alcohol reduction apps on the UK Apple app store and aims to help hazardous and harmful drinkers reduce their alcohol consumption. *Drink Less* is capable of reaching a large proportion of the UK population at a low incremental cost. The development and evaluation of *Drink Less* was guided by the Medical Research Council’s guidance on complex interventions [15] and the Multiphase Optimisation Strategy [16]. The development of the *Drink Less* app was informed by the COM-B model of behaviour [17] and multiple sources of evidence [14,18,19]; and is reported in full elsewhere [12]. A factorial screening trial was conducted to identify the most effective modules (distinct behaviour change interventions) within the app, which established that four of the five modules appeared to have an effect on reducing alcohol consumption after four weeks if combined with one of the other modules [13]. Data also suggested that users of *Drink Less* found it to be engaging [13], which is important to reduce participant attrition.

The strategy for refining the effectiveness and usability of the app was based on: i) findings from the previous factorial trial, ii) a content analysis of user feedback, and iii) an updated evidence review and meta-analysis of behaviour change techniques in digital alcohol interventions. The refinement process is reported in full elsewhere [*in preparation*]. Both the initial development and refinement of *Drink Less* have involved input from users across the social spectrum on the functionality, design, and language used in the app [20]. The same approach was used for a smoking cessation digital intervention that was found to be effective for increasing smoking cessation rates across the social spectrum [21].

The next step in the Multiphase Optimisation Strategy is to conduct a RCT to evaluate the long-term effectiveness and cost-effectiveness of the digital recommendation of the refined *Drink Less* app, compared with alcohol advice from the NHS alcohol advice webpage (usual digital care, available to anyone seeking alcohol support), in reducing alcohol consumption among hazardous and harmful drinkers. This research will be the first RCT of an alcohol reduction app for the general population in the UK and will evaluate whether it is worth investing resources into promoting and disseminating the app on a larger scale.

### Research questions

1. At a 6-month follow-up, does the digital recommendation to use *Drink Less* compared with the NHS alcohol advice webpage to hazardous and harmful drinkers:
  a. Reduce weekly alcohol consumption (in UK standard units)?
  b. Reduce heavy episodic alcohol consumption?
  c. Reduce the proportion of hazardous drinkers?
  d. Reduce alcohol-related problems and injury, and use of healthcare services?
  e. Improve health-related quality of life?
2. What is the extent of user engagement with *Drink Less* and does user engagement moderate these outcomes?
3. Through what psychological measures does engagement with *Drink Less* change drinking behaviour?
4. What are participants’ views on the acceptability of the intervention?
5. What is the cost-utility and potential impact on health inequalities of *Drink Less* compared with the NHS alcohol advice webpage in terms of reduction in alcohol consumption and health-related quality of life using a short time horizon?
6. What is the longer-term cost-effectiveness and potential impact on health inequalities of *Drink Less* compared with the NHS alcohol advice webpage, if rolled out on a national level through active promotion to the public, over a 20-year period?

## Methods

### Design

A two-arm, parallel group, RCT with a 1:1 allocation comparing the intervention (*Drink Less*) with usual digital care (the NHS alcohol advice webpage), with an embedded mixed-methods process evaluation.

### Setting

The study will take place online with participants who live in the UK.

### Participants

Participants will be included if they: are aged 18 years or over, live in the UK, are hazardous and harmful drinkers (Alcohol Use Disorders Identification Test (AUDIT) score>=8), have access to an iOS device (i.e. iPhone, iPod touch or iPad), and want to drink less alcohol. Participants will be excluded if they are unwilling to complete follow-up assessments or are unable to read English (for pragmatic reasons).

### Recruitment

Recruitment is due to run from July 2020 to March 2022 via a multi-pronged strategy including: an advertisement on the NHS website; a mail-out to a database of UK-based users of the Smoke Free app; and press releases and local advertising through health care providers and/or national and local government colleagues. The advertisements will be co-developed with public representatives.

### Sample size

A sample size of 5562 participants (2781 in the comparator group and 2781 in the intervention group) is required to detect a mean difference reduction of 2 UK units (16g of alcohol) in last week alcohol consumption at 90% power with an alpha of 0.05 and a two-tailed test. This was calculated using G*Power software [22]. The estimated effect size is in line with the Cochrane review on digital alcohol interventions [5] and is roughly equivalent to that found in face-to-face brief interventions [23].

A sub-sample of 26 participants (13 from each group [24]), who consented to a short interview about their experience of the trial, will be selected after the 6-month follow-up, as part of the mixed-methods process evaluation. Participants will be purposively sampled to achieve good diversity in terms of socio-demographic characteristics, and with high and low engagement. Data will be analysed and data collection will continue in an iterative process (adding 10 participants at a time) until thematic ‘meaning’ saturation is reached (see Analysis).

### Intervention

*Drink Less* is a stand-alone app-based intervention that is freely available via the Apple app store in the UK [25]. *Drink Less* was developed for hazardous and harmful drinkers to help them reduce their alcohol consumption. *Drink Less* consists of evidence-based modules to help users change their drinking behaviour: Goal Setting; Self-monitoring & Feedback; Action Planning; Normative Feedback, Cognitive Bias Re-training, Behavioural Substitution and Information about Antecedents, which map to behaviour change techniques (see Figure 1). The development and content of the original *Drink Less* version is reported in full elsewhere [12] and the refined version is reported online (https://osf.io/mc8yz/). The app contains standard features such as the UK Chief Medical Officers’ low-risk drinking guidelines (14 units a week) [26].

**Figure 1:**
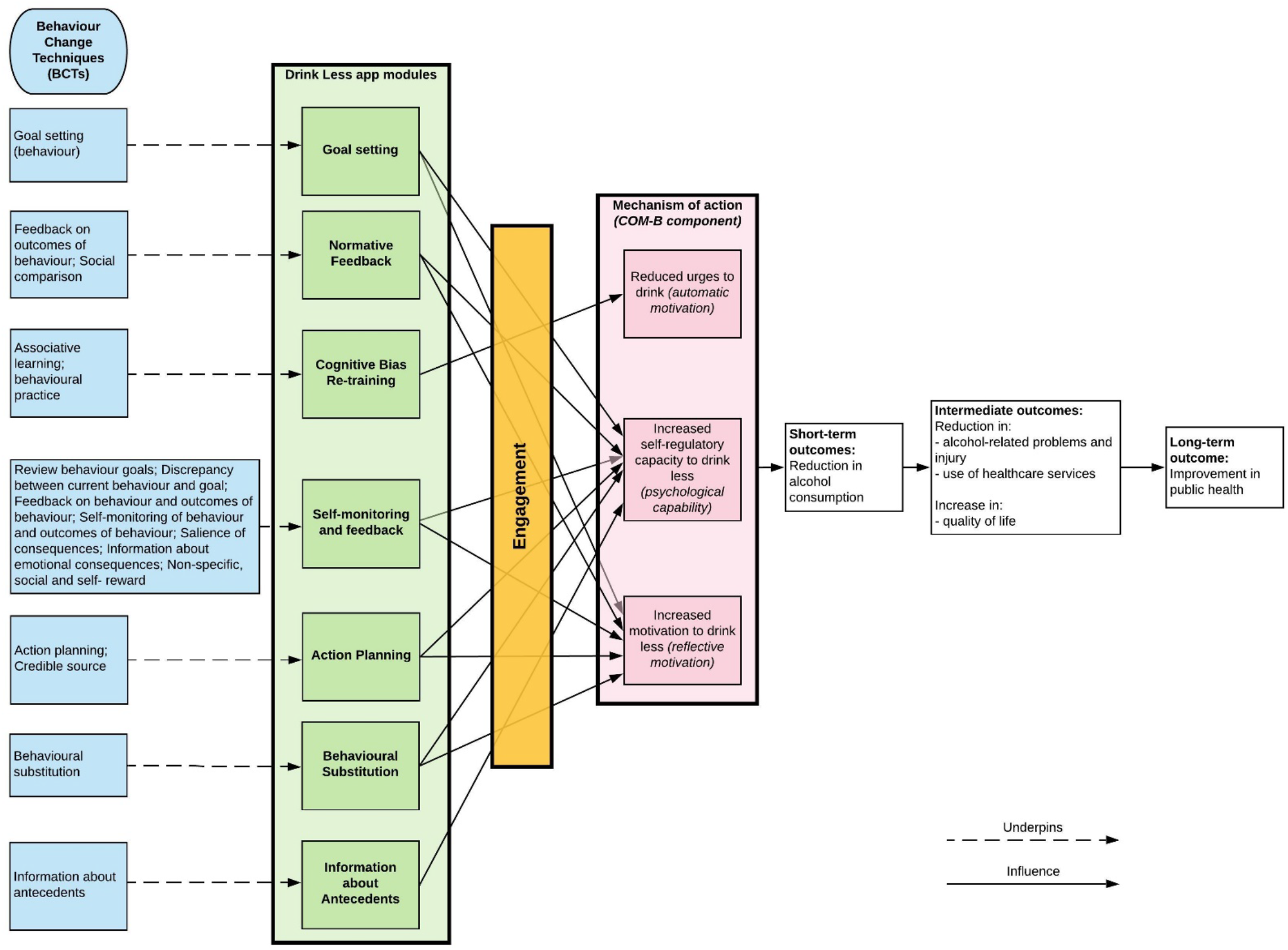
Logic model for the process of change of the Drink Less app (developed by the internal research team)

On downloading the app, users are asked to complete the AUDIT, provide socio-demographic details and then receive the Normative Feedback module. Users are then guided through setting a goal and shown how to use the key features of the app. Users can access all of the modules from the dashboard and the menu bar. The dashboard (the landing page of the app) has suggestions for the user to complete each day, as well as features of and links to the modules. Users can choose to have daily reminders to complete their drinks and mood diary for the previous day. The app provides a ‘toolbox’ of features for users to choose from and access as and when they want. The app is not tailored to the user except for personalised feedback in two modules: Normative Feedback and Self-monitoring & Feedback. Any modifications to the app during the trial (e.g., bug fixes) will be documented and reported.

*Drink Less* is expected to reduce the alcohol consumption of its users based on (i) its robust theoretical basis (the COM-B model of behaviour [17]), (ii) its evidence base, and (iii) user feedback that indicates users believe that it helps them to reduce their drinking and has a positive effect on their health and well-being. It is also expected to reduce urges to drink, increase motivation to drink less, and increase self-regulatory capacity to drink less (see Figure 1 for the logic model). *Drink Less* is also highly rated by users (average 4.2-star rating in the Apple (UK) App Store with over 60,000 unique users as of 1^st^ November 2019).

### Comparator

The comparator group will receive the recommendation to view the NHS alcohol advice webpage [27]. The NHS webpage also has links to other webpages aimed at hazardous and harmful drinkers (e.g., ‘Tips on cutting down’ and ‘Benefits of cutting down’). This can be considered reflective of ‘usual digital care’ in this context as it is the digital support currently available to treatment-seeking individuals from the NHS. Therefore, this comparator best serves the primary purpose of the trial [28] which is to investigate whether it is worth promoting *Drink Less* over the ‘usual digital care’, and is of direct policy relevance. Furthermore, it is important to have a comparator that is relevant to the same target population as the intervention, and both *Drink Less* and the NHS webpage are aimed at adults in the general population. Any changes to the comparator during the trial will be documented.

### Procedure

Figure 2 illustrates the study design and flow of participants and Tables and figures Table 1 summarises the schedule of enrolment and follow-up assessment for trial participants.

**Table 1:**
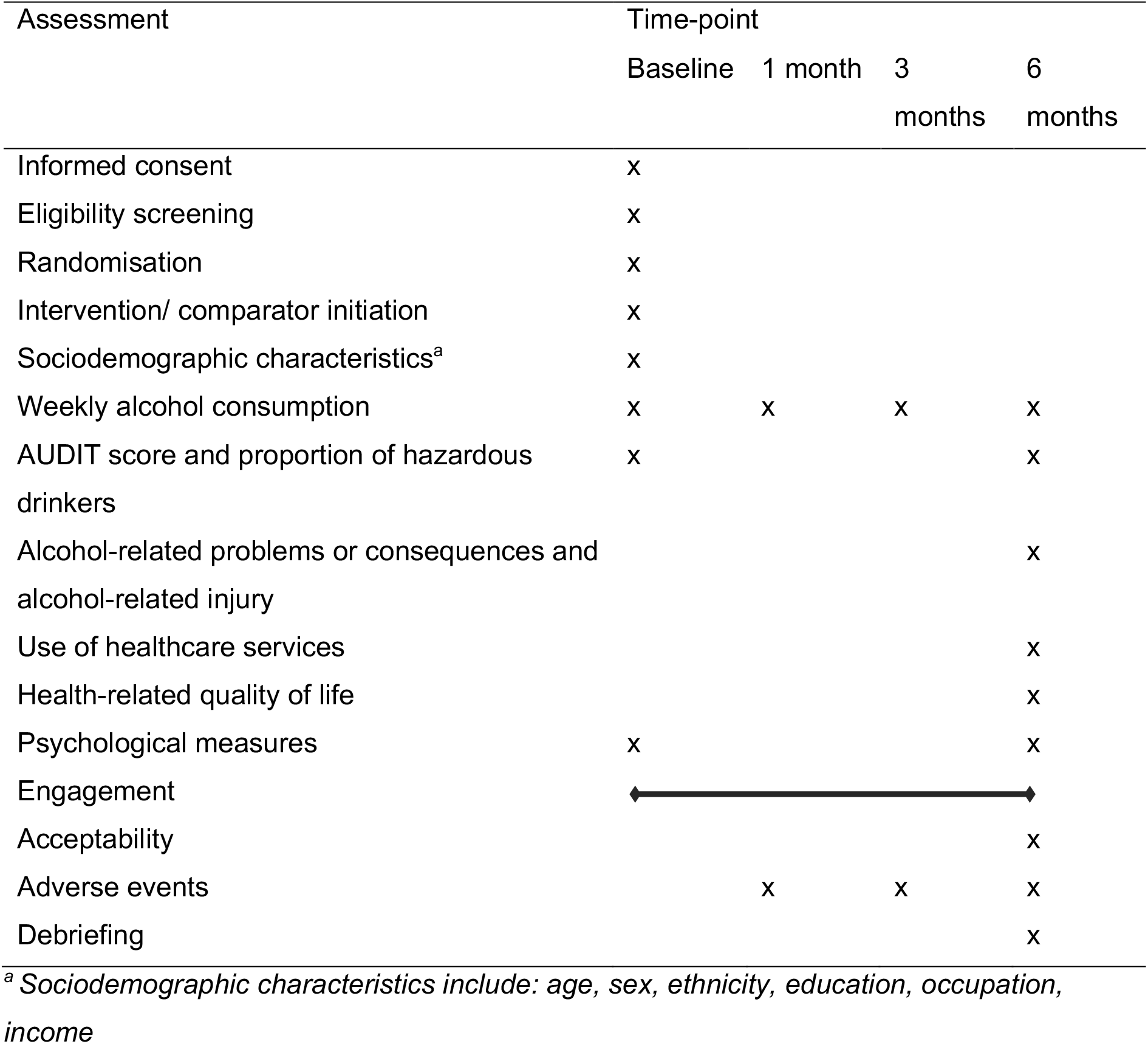
Schedule of enrolment and follow-up assessments.

**Figure 2:**
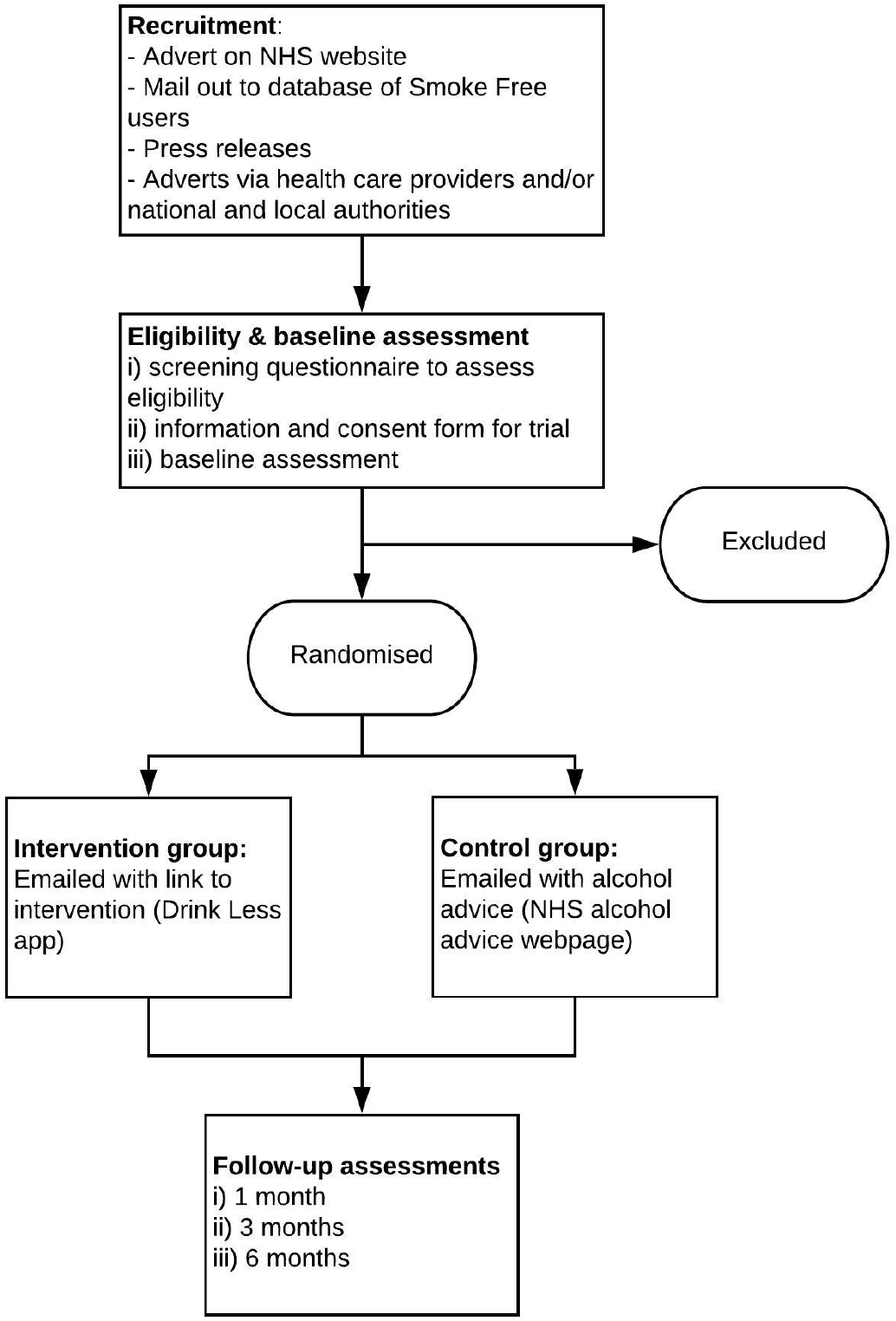
Flowchart of procedure.

#### Eligibility Assessment

Participants will self-enrol into the study and potential participants will be asked to respond to a web-based screening questionnaire to assess the inclusion and exclusion criteria.

#### Consenting

If people meet the eligibility criteria they will be shown the information sheet [*link here*] and informed that they will be re-contacted on three occasions (at 1, 3 and 6 months). Participants will then be asked to provide consent online to participate in the study.

#### Baseline Assessment

Participants will complete a web-based assessment of socio-demographic measures and the AUDIT (see Measures), and their contact details (email address, telephone number and postal address) for follow-up assessments.

#### Randomisation

Participants who complete the baseline assessment will be randomised individually to intervention and comparator groups using block randomisation (block size of 50) and a random allocation sequence generated by an online automated algorithm (at a ratio of 1:1). Participants will be blinded to study arm. There will be no involvement of the researchers in the randomisation process and there will be complete allocation concealment.

#### Intervention and Comparator Delivery

Participants will be emailed within 24 hours of the baseline assessment and randomisation with the recommendation to either use *Drink Less* (the intervention) or the NHS alcohol advice webpage (comparator). Participants allocated to the intervention condition will be provided with instructions on how to download the *Drink Less* app along with the contact details of the project team who will provide ongoing technical support. These emails will be co-developed with public representatives.

#### Follow-Up Assessments

Follow-up assessments will be conducted 1, 3 and 6 months after baseline. The 6-month follow-up assessment will assess the primary and secondary outcome measures, and psychological measures; the follow-up assessments at 1 and 3 months will assess the primary outcome measure only. Participants will have up to 30 days to complete each survey to maximise data retention. Participants will be sent up to three automated emails with a link to a web-based survey for the follow-up assessments on days 0, 5 and 9. Participants who do not complete the web-based follow-up assessment will be sequentially offered opportunities to do so via phone (called once per day on days 10-17), mailed survey (on day 18) and by postcard (on day 30). Participants will be compensated with gift vouchers of up to £36 for completing the three surveys: £6 for the survey at 1 and 3 months; £12 at 6 months with an additional £12 if the 6-month survey is completed within 24 hours.

At the six-month follow, participants will indicate whether they are happy to be called for a short interview about their experience of the trial; there is no additional compensation for this telephone interview. Acceptability will then be measured after the 6-month follow-up via telephone interviews.

Participants will be asked whether they experienced any unexpected consequences, adverse events or other harms from participating in the study (in an open-ended question at the 1-, 3- and 6-month follow-up), and whether they have used any other forms of support for alcohol reduction (at the 6-month follow-up).

#### Debriefing

On completion of the trial, after the final follow-up at 6 months, the comparator group will also be informed about the intervention, *Drink Less*.

### Measures

#### Sociodemographic measures

Sociodemographic measures will be assessed at baseline: age (in years, continuous), sex (% female), ethnicity (% white), education (% post-16 educational qualifications), occupation (to derive social grade AB, C1, C2, D, E dichotomised into: ABC1 vs. C2DE), and annual household income (% >£26,000 [29]).

#### COVID-19 measures

The recent COVID-19 pandemic is affecting many aspects of people’s lives in the UK. A lockdown is currently in place, limiting many people’s ability to leave their homes, apart from essential journeys. Early evidence suggests that COVID-19 may affect alcohol consumption, some lighter drinkers are drinking less than usual and some heavier drinkers are drinking more. In order to assess for the effects of the pandemic in the analysis, participants will respond to a brief COVID-19 survey at each time point. Users will be asked “Do you currently feel like COVID-19 is affecting your alcohol consumption and how you feel about drinking alcohol?” If participants respond ‘no’ they will continue with the rest of the survey. Participants responding “yes” will be asked to answer five follow up questions assessing the extent to which the pandemic is affecting their concerns about their alcohol consumption, their motivation to cut down and their patterns of consumption. Change in concerns about drinking will be measured by the question “Is COVID-19 and its associated effects (e.g. financial, social or health) currently affecting how worried you feel about your alcohol consumption?” followed by three response options ‘more worried’, ‘no change’ and ‘less worried’. Change in motivation to reduce alcohol consumption will be measured by the question “Is COVID-19 and its associated effects currently affecting your motivation to reduce your alcohol consumption?” with three response options ‘more motivated’, ‘no change’ and ‘less motivated’. Three questions measure changes in drinking patterns. Change in the frequency of drinking is measured by the question “Is COVID-19 and its associated effects affecting how frequently you consume alcohol?” with three response options ‘consume alcohol more frequently’, ‘no change’ and ‘consume alcohol less frequently’. Change in the volume of alcohol consumed is measured by the question “Is COVID-19 and its associated effects currently affecting how many units of alcohol you generally consume when you do drink?” with three response options ‘generally drink more units’, ‘no change’ and ‘generally drink less units’. Finally, change in the frequency of binge drinking is measured by the question “Is COVID-19 and its associated effects currently affecting how often you consume 6 or more units of alcohol on a single occasion?” with three response options ‘more likely to consume 6 or more units on a single occasion’, ‘no change’, and ‘less likely to consume 6 or more units on a single occasion’. The date of national responses to COVID-19 (e.g. lockdown) will also be monitored and recorded by the research team.

#### Primary outcome measure

The primary outcome measure is change between baseline and 6-month follow-up in self-reported weekly alcohol consumption estimated over the last month, in UK standard units. Change in weekly alcohol consumption will be derived from the extended quantity-frequency questions of the AUDIT [30], adjusting for heavy episodic use (question 3 of the AUDIT) and therefore allocating an individual to a category of consumption that is closest to their actual consumption. The extended quantity-frequency questions of AUDIT exhibit similar sensitivity and specificity to the full AUDIT [31] and have demonstrated excellent reliability and responsiveness to short-term change [32]. This method of deriving alcohol consumption has been used in other trials [33–36] and has high levels of agreement in levels of self-reported consumption when compared with other retrospective daily diary measures [37,38]. This measure minimises response burden on participants due to its brevity, which is a critical issue in digital trials that have minimal contact with participants and can suffer from high levels of attrition [39].

#### Secondary outcome measures

- Change between baseline and 1- and 3-month follow-ups in self-reported weekly alcohol consumption estimated over the last month;
- Heavy episodic alcohol use (measured using AUDIT question 3) at 6-month follow-up
- Proportion of hazardous drinkers (AUDIT score>=8);
- Alcohol-related problems or consequences and alcohol-related injury (measured using the Alcohol Short Index of Problems [40]) at 6-month follow-up;
- Use of healthcare services (measured using the Service Use Questionnaire [41,42]) at 6-month follow-up;
- Health-related quality of life (measured using the EQ-5D-5L) at 6-month follow-up;

The self-report AUDIT questionnaire has 10-items that measure alcohol consumption, harms and dependence. The AUDIT is a reliable and standardised alcohol-related outcome measure that is commonly used in alcohol trials [5,23], has high test-retest reliability when completed online [43], and allows the derivation of a core outcome set for consistency across trials, and to minimise research waste and selective reporting [44].

#### Process measures

The mixed-methods process evaluation involves assessing psychological measures, engagement and acceptability.

Psychological measures will be assessed as potential mechanisms of action at baseline and 6-month follow-up using four theoretical measures: urges to drink; motivation to drink less; self-regulatory and self-monitoring capacity (see Figure 1 for the Logic Model). Strength of urges to drink will be measured by the question “How strongly have you felt the urge to drink alcohol in the past 24 hours?” with six options from ‘Not at all’ to ‘Extremely strong’. The strength of urges to drink measure has been chosen as it can be done through a single item question and has fair long-term test re-test reliability [45]. The motivation to drink less will be measured with the single-item Motivation to Stop Scale (MTSS). The MTSS and the urges to drink measure are both used in the Alcohol Toolkit Study allowing for national comparisons and have been successfully used in an observational study that estimates patterns of alcohol consumption and reduction in a sample in England [46]. Self-regulatory capacity will be measured by “How difficult do you find it to control your drinking?” using a 5-point scale from ‘Not at all’ to ‘Extremely’. Self-monitoring capacity will be measured by “How often, if at all, do you keep track of how many units of alcohol you personally drink each week?” ranging from ‘Never’ to ‘Always’.

Engagement with *Drink Less* will be assessed in terms of app download, and frequency, amount, duration, and depth of engagement [47] – all automatically recorded within the app, among participants in the intervention group. This will provide objective data on how participants interact with the app. App download will be assessed by whether the participant downloaded and opened *Drink Less* (a binary yes/no measure). Frequency of engagement will be assessed by number of sessions, where a new session is defined as a new screen view after 30 minutes of inactivity [48]. Amount of engagement will be assessed by time on app, in minutes. Duration of engagement will be assessed by number of days used. Depth of engagement will be assessed by the percentage of available screens viewed. Participants will also be asked about their use of the app in the semi-structured interviews to complement and enhance the patterns identified from the objective engagement data. For example: “Can you tell me about your experience of using the app?”, and “In what situations did you use it and why?” Adherence to either the intervention or comparator will be measured as to whether the link in the email to download *Drink Less* or view the NHS webpage, respectively, was clicked on via a mailing system.

The acceptability of the intervention will be assessed in a number of short semi-structured interviews among a sub-sample of participants in both the intervention and comparator group after the 6 month follow-up. The interview will focus on perceptions of the intervention in terms of the acceptability of the app – the extent to which participants consider the app to be appropriate, based on anticipated or experienced cognitive and emotional responses to the intervention [49]. The interview topic guide will be based on the Theoretical Framework of Acceptability [49].

#### Economic measures

Unit costs for the economic evaluation will be taken from standard sources (e.g., PSSRU, NHS tariffs).

### Data management and monitoring

Baseline and follow-up assessment data will be collected online and held securely in Data Safe Haven. All personal data will be pseudonymised. Engagement data will be collected automatically from the app and downloaded via python/pandas script into Data Safe Haven from a secure https protocol and ‘Nodechef’ (an online platform for hosting mobile apps). The audio recording of the semi-structured interviews on acceptability will be pseudonymised and transferred directly from the recording device to Data Safe Haven. Any participant who opts out of the study will have their data deleted.

An independent data monitoring committee will have access to the unblinded comparative data and will monitor these data. The committee will make recommendations on whether there are any reasons to terminate the trial that the chair will report to the independent trial steering committee.

### Analysis

The data will be analysed using R Studio [50]. The data analyst will be blinded to participants’ group and the analysis plan will be finalised and uploaded onto Open Science Framework prior to the start of data analysis when the trial will be analysed in accordance with the pre-specified plan.

Descriptive statistics of participants’ sociodemographic characteristics and AUDIT score will be reported for who the study recruited and who then accessed the intervention. The difference between the intervention and comparator groups on baseline characteristics will be assessed using one-way ANOVAs for continuous variables (age, AUDIT score) and 2-sided chi-squared tests (or Fisher’s exact test for rare events) for categorical variables (sex, ethnicity, education, occupation, income, COVID-19 survey measures).

ANOVAs are generally considered robust against small deviations from the normality assumption with only a small effect on the Type I error rate [51]. However, if there is evidence of significant deviation we will attempt to resolve this with transformations (e.g., logarithmic or square root transformations) or choose the nonparametric Kruskal-Wallis H Test which does not require the assumption of normality.

#### Analysis of primary and secondary outcomes

The primary analysis will use a conservative intention-to-treat approach to missing data with the assumption of no change for participants who do not respond to follow-up (i.e., analysis of outcome data from all randomised participants). The effect of group allocation on the primary outcome, change in weekly alcohol consumption, will be examined with a one-way ANCOVA, adjusting for baseline consumption [52–54].

A secondary analysis will be conducted to assess differences in change in weekly alcohol consumption using a one-way ANOVA. ANCOVA and ANOVA have several statistical assumptions which will be assessed (i.e., the errors of the data are normally distributed and there is equal variances between treatments). The analyses are robust to slight deviations in parametric assumptions, but where large deviations are found data transformations and appropriate non-parametric tests will be considered.

Additional sensitivity analyses will be conducted for the primary outcome at 6 months: 1) responders-only (i.e. those who completed the 6-month follow-up survey); 2) using multiple imputation for non-responders on baseline characteristics (with five imputed data sets [55] combined using Rubin’s rules [56]) and assuming a normal distribution with a mean of 0 and SD reflecting the variation in change among responders; 3) per-protocol approach whereby only participants who downloaded *Drink Less* in the intervention group and those who clicked on the link to the NHS alcohol advice webpage in the comparator group are included in the analyses, and whereby participants whose treatment was contaminated are excluded; 4) an instrument variable analysis accounting for non-use in the intervention group and contamination in the comparator by operationalising the difference in app usage between the two conditions, and 5) last observation carried forward.

Secondary analyses will assess: 1) the secondary outcomes at 6 months using ANOVA and chi-squared analyses as appropriate and 2) the change over time in the primary outcome using 1- and 3-month follow-up data.

Confidence intervals, effect sizes (partial eta squared for ANOVA analyses, odds ratios for chi-squared and regression analyses), and exact p values will be reported. Bayes Factors will be calculated using a half normal distribution to specify the predicted effect (of a 2 UK unit reduction per week) with a peak at 0 (no effect) and the standard deviation equal to the expected effect size with Robustness Regions reported to specify the range of expected effect sizes that support the same conclusion [57].

Finally, interactions will be assessed between group allocation with age, sex, ethnicity, education, occupation, income, and COVID-19 measures (survey and national responses) for primary and secondary outcomes. Where significant interactions are found the findings will be stratified, drawing on the PROGRESS-Plus framework to explicitly consider health equity between the intervention and comparator group [58].

#### Process evaluation

The process evaluation will involve quantitative analysis of the psychological and engagement measures and qualitative analysis of interview transcripts relating to the acceptability of the intervention.

The extent of user engagement with *Drink Less* will be evaluated through descriptive statistics of the engagement measures. Detailed modelling of variations between participants will be conducted to explore variation in engagement and psychological measures by sociodemographic characteristics, baseline AUDIT scores and COVID-19 measures (survey and national responses). A mediation analysis will be conducted to determine if any effect of group allocation on the primary outcome is mediated by changes in the psychological measures. The psychological measures will also be integrated into the modelling of effectiveness outcomes for *Drink Less* to identify links between the outcomes, participant engagement and psychological measures.

Anonymised interview transcripts on the intervention’s acceptability will be analysed using a combined framework and thematic analysis approach. This involves initially coding participant responses according to the TFA construct they are judged to represent best, then grouping similar responses within each construct inductively to generate content themes representing how that construct contributes to reported acceptability. Twenty-six participants will be interviewed initially (13 from each group [24]), then data will be analysed and data collection will continue in an iterative process (adding 10 participants at a time) until thematic ‘meaning’ saturation is reached. Meaning saturation is defined as the point at which the issues are fully understood and no further dimensions, nuances, or insights are found [59].

#### Health economic evaluation

The economic evaluation will take a two-stage approach to analyse the cost-utility of *Drink Less* from the NHS perspective. The first stage will be an analysis of the cost-effectiveness of the app in the trial population over the duration of the trial itself (including follow-up). Costs will include the cost of the interventions in both arms and the cost of NHS resource use (i.e., cost of changes in service use and treatments). The cost-effectiveness analysis will take into account the total development cost of *Drink Less* but keep it separate from the incremental evaluation as there are no anticipated additional costs per user using the app. Effects will be measured in terms of i) reduction in alcohol consumption and ii) health-related quality of life, measured in Quality-Adjusted Life Years (QALYs). The cost-utility will be measured in terms of Incremental Cost-Effectiveness Ratio, the ratio between the difference in costs and difference in effects between the intervention and comparator groups.

As there can be a delay of several years between reductions in alcohol consumption and improvements in health [60], the full impacts of interventions designed to reduce alcohol consumption on health and healthcare costs may not be seen until well beyond the time horizon of an RCT. The second stage of the economic evaluation will address this limitation by using the established and widely-used Sheffield Alcohol Policy Model [61,62] to assess the longer-term cost-effectiveness of the intervention, if rolled out on a national level through active promotion to the public, over a 20-year time horizon.

Both short- and long-term evaluations will assess the impact on health inequalities using a Distribution Cost-Effectiveness Analysis framework [63,64]. Costs and QALY outcomes will be estimated separately by socioeconomic group, defined by social grade (AB, C1, C2, D or E). These group-specific results will be combined with estimates of health pre-intervention and the opportunity cost of additional healthcare spending to place the intervention on the ‘health equity impact plane’. Published estimates of inequality aversion, quantifying the extent to which society is willing to trade off changes in cost-effectiveness for changes in health inequalities, will be used to identify the optimal strategy after accounting for the inequality impacts of each approach.

### Ethical approval

Ethical approval has been obtained from UCL Research Ethics Committee (16799/001).

### Dissemination policy

Results will be disseminated by open-access peer-reviewed journal articles, presentations at scientific conferences, press releases, a stakeholder workshop, and blog posts. NIHR authorship guidelines will be followed. Study materials, anonymised data and code will be made available on Open Science Framework (on the project page: osf.io/q8mua), and the source code for the app will be released under the GNU General Public License (v3) on Github.

## Discussion

### Key outputs

This study will provide evidence on the effectiveness and cost-effectiveness of the Drink Less app – a theory- and evidence-informed intervention – for the reduction of hazardous and harmful alcohol consumption. The embedded mixed-methods process evaluation will evaluate the extent of user engagement with *Drink Less*, and whether user engagement moderates or changes in psychological measures mediate the effectiveness of the intervention and participants’ views on the acceptability of the intervention.

### Implications

The use of behavioural science in the development and refinement of the *Drink Less* app and the trial methodology will improve the understanding of what works for whom and why. *Drink Less* collects detailed user engagement and behavioural data of potential value in advancing its underpinning theory [65], which can inform future behaviour change interventions. The principles of Open Science will continue to be followed, which is important for efficient scientific progress.

*Drink Less* is currently only available on iOS devices though this RCT will provide a proof of concept and strong rationale, if effective, for developing a native Android version, which would maximise the potential impact of *Drink* Less on public health.

*Drink Less* is in a good position for implementation in healthcare settings as it is already aligned with the criteria from the NHS Digital Assessment Questionnaire and with the NICE Evidence for Effectiveness Framework and Code of Conduct for Digital Health [66–68]. The app will be submitted to the NHS Apps Library if found to be effective, which may result in healthcare practitioners recommending or prescribing the app to patients [69]. *Drink Less* is a standalone app and therefore in a strong position to be actively promoted on a national level, through media contacts and relevant organisations, to encourage use of the app amongst wider society. A sustainability model will be developed as part of this research project to maximise the impact of the app by enabling long-term planning for its implementation, adoption and future development, and will inform the long-term economic analysis in terms of potential reach and costs.

### Anticipated challenges

The two main anticipated challenges are recruitment and retention rates. A multi-pronged recruitment strategy is planned, including placing an advert on a NHS webpage, which provides the ability to access a large number of drinkers. A higher rate of recruitment than is currently required (265 participants per month) was achieved in the factorial trial of *Drink Less* (355 per month) [13] with web-based recruitment and screening. However, a contingency plan is in place that involves advertisements placed on Google and Facebook (following the methods used in a similar trial [70]) and use of a recruitment company. We will also calculate Bayes factors, which in the situation of under recruitment will allow us to distinguish between two interpretations of a non-significant result: i) support for the null hypothesis of ‘no effect’ and ii) data are insensitive to detect an effect [71,72]. To maximise the overall retention rate, financial incentives will be offered as a reasonable acknowledgement of participants’ time, and a range of methods will be used for follow-up contact sequentially, which has been shown to improve response rates [73]. The trial cost per participant and methods of follow-up contact are comparable with similar studies that achieved high retention rates [70,74]. The intention-to-treat approach to analysis is conservative, meaning that if a follow-up rate similar to that in other trials is achieved, then if anything the effect size will be an underestimate.

We acknowledge that the research team is evaluating an app that it has developed and that there may be a perceived bias. The team have no financial stake in the app and the code is available in Github by the GNU General Public License (v3). The trial has been designed to meet the Cochrane Risk of Bias criterion of ‘low risk’ and we have followed best practice guidelines to ensure a robust evaluation is conducted, including blinding and oversight by independent trial steering and data monitoring committees. The data will be analysed in accordance with a pre-specified analysis plan approved by the committees and uploaded onto Open Science Framework prior to the start of data analysis.

## Conclusions

This study will be the first RCT of an alcohol reduction app for the general population in the UK, thereby starting to build a strong evidence base on the effectiveness and cost-effectiveness of health-related apps. The RCT will be able to directly inform the pragmatic question of interest: whether it is worth investing resources into promoting the app.

## Data Availability

Study materials, anonymised data and code will be made available on Open Science Framework (on the project page: osf.io/q8mua).

## Acknowledgements

This project is funded by the National Institute for Health Research (NIHR) [Public Health Research Programme (project reference NIHR127651)]. The views expressed are those of the author(s) and not necessarily those of the National Institute for Health Research or the Department of Health and Social Care, or Public Health England (PHE).

CG, EB and JB are also funded by Cancer Research UK (CRUK: C1417/A22962). Drink Less was funded by the NIHR School for Public Health Research (SPHR), the UK Centre for Tobacco and Alcohol Studies (UKCTAS), the Society for the Study of Addiction (SSA) and CRUK. EP is supported by the NIHR ARC North Thames. MH acknowledges NIHR Health Protection Research Unit in Evaluation, and MM and MH acknowledge NIHR Biomedical Research Centre at Bristol.

JB and MM are part of the SPECTRUM Consortium, UK.

The funders played no role in the design, conduct or analysis of the study, nor in the interpretation or reporting of study findings.

We would like to thank Dr Dave Crane for his important role in the development and factorial screening trial of the Drink Less app.

